# Longitudinal SARS-CoV-2 serosurveillance of over ten thousand health care workers in the Providence Oregon cohort

**DOI:** 10.1101/2020.08.16.20176107

**Authors:** Rom Leidner, Angi Frary, Julie Cramer, David Ball, Roshanthi Weerasinghe, Mark Schmidt, Justin Jin, Veronica Luzzi, Alec Saitman, Jeffrey A. Young, David Leidner, Kendall Sawa, Scott Marsal, Kevin Olson, Nancy Frisco, Amy Compton-Phillips, Walter Urba, Brian Piening, Carlo Bifulco

## Abstract

Frontline healthcare workers (HCW) are a high-risk population for SARS-CoV-2 infection. Here we present results from a large serosurveillance study of 10,019 asymptomatic HCW conducted during April-May 2020, in eight hospital medical centers across the state of Oregon, USA during the initial peak of the pandemic. Free and voluntary testing was performed at 14 +/− 3 day intervals, over a 4-week window at each site, utilizing a lab-developed ELISA based on the Epitope Diagnostics COVID-19 nucleocapsid IgG detection Kit. We identified 253 SARS-CoV-2 IgG seropositive individuals among 10,019 total participants, representing a cross-sectional seroprevalence of 2.53%. Subgroup analysis identified differential seropositivity by job role, ranging from 8.03% among housekeepers, odds ratio 3.17 (95% CI 1.59–5.71), to 0.00% among anesthesiologists, odds ratio 0.00 (95% CI 0–0.26), both of which were significant. Over the course of the study, 17 seroconversions (0.25%) and 101 seroreversions (1.50%) were identified. Self-reported SARS-CoV-2 swab qPCR testing, when compared with subsequent serology on study, showed only modest agreement, **κ** = 0.47 (95% CI 0.32–0.62). Overall, these findings demonstrate relatively low seroprevalence and very low seroconversion rates among HCW in Oregon, USA, over a period in which aggressive social distancing measures were in place. The high rate of seroreversion observed in this cohort, and the relatively high discordance between SARS-CoV-2 serology and swab qPCR, highlight limitations of current detection methods, and stress the need for development of novel assessment methodologies to more accurately identify exposure (and/or immunity) to SARS-CoV-2 in this population.

## INTRODUCTION

The original outbreak of COVID-19 in Wuhan, China, caused by SARS-CoV-2, a novel strain of betacoronavirus that is capable of person-to-person spread, has rapidly become a global pandemic^1^. The crisis has been hallmarked by high transmission rates leading to a rapid increase in critically ill patients that continue to overwhelm intensive care units in multiple communities throughout the world^2^. To combat the spread of the virus, governments have taken steps such as travel restrictions, mandatory utilization of personal protective equipment (PPE), sheltering in place and quarantines^3^. Healthcare workers (HCWs) providing direct care for COVID-19 patients are at a significant risk of infection and have accounted for a significant proportion of COVID19 cases^4–6^. HCWs at Providence St. Joseph Health (PSJH), a large integrated not-for-profit health care system operating in seven US states, have been at the forefront of the crisis since the first reported U.S. COVID-19 patient was admitted to Providence Regional Medical Center Everett in January 2020^7^. “Serosurvey” efforts, aimed at determining overall exposure and seroconversion rates in a population, provide a tool to identify exposure associated risk factors and can guide the rational utilization in a health care setting of personal protective equipment (PPE), in addition to return to work policies and providing psychological support to health care workers who are at risk of secondary mental health issues due to overwork and fear of contagion^8^. Although serological studies have been extensively used to determine local epidemic dynamics and population infection fatality rates, significant gaps persist in the current understanding of the humoral responses to SARS-CoV-2 in individuals with asymptomatic infections, with recent evidence suggesting that asymptomatic individuals may have a weaker humoral immune response to SARS-CoV-2 infection and an early reduction in antibody levels^9–13^.

## METHODS

Under an IRB approved protocol (PSJH IRB2020000221, entitled: SARS-CoV-2 Serosurveillance Protocol for PSJH Health Care Workers), between April 8 and May 22, SARS-CoV-2 nucleocapsid IgG antibody serosurveillance was offered to asymptomatic HCW at 8 Oregon USA Providence medical centers, (Fig. 1). Testing was free and voluntary, with confidential results notification, and was offered to all HCW with direct patient contact, contact with patient biospecimens or patient linens. Testing was performed every 14 +/− 3 days over a four week interval at each site. The opening of each site was slightly staggered, with initial activation in the two largest Portland, Oregon city hospitals. Specific dates of testing are provided in Table 1. Test results were accessed by participants via an auto-generated e-mail link to an intranet hosted secure web application used to display results and dynamically updated institutional guidelines. Participants were asked to provide basic demographic information including age, gender, job role (one only), workplace setting (up to three, e.g. x-ray tech in ER, ICU and wards), previous swab PCR testing for SARS-CoV-2, and if so, whether positive (dates not queried). Results reporting was confidential and not shared with the employer or unit managers. Residual sera were cryopreserved for additional analyses. A dedicated study e-mail inbox for participant questions was continuously staffed and received roughly 2,700 queries over the course of the study, indicative of HCW uncertainty/anxiety levels in April 2020.

**Table 1.** Serosurveillance Providence Oregon Cohort: Asymptomatic Health Care Workers

| Providence Medical Center | Serosurveillance every 14 +/- 3 days | Test cycle 1 Pos/Total (%) | Test cycle 2 Pos/Total | Test cycle 3 Pos/Total |
| --- | --- | --- | --- | --- |
| Portland | April 8 - May 8 | +94/3,452 (2.72%) | +46/2,612 | +17/797 |
| St. Vincent | April 13 - May 8 | +79/3,251 (2.43%) | +64/2,140 | +9/91 |
| Milwaukie | April 16 - May 14 | +17/560 (3.04%) | +11/372 | +2/99 |
| Newberg | April 20 - May 15 | +14/603 (2.32%) | +7/319 | +0/12 |
| Willamette Falls | April 17 - May 15 | +10/660 (1.52%) | +5/420 | +0/62 |
| Hood River | April 27 - May 22 | +5/441 (1.13%) | +2/115 | +0/4 |
| Medford | April 20 - May 8 | +12/760 (1.58%) | +6/586 | +0/0 |
| Seaside | April 22 - May 20 | +5/285 (1.75%) | +5/153 | +0/7 |
| **Totals** | **2.53% (+253/10,019)** | **+236/10,012<sup>a</sup> (2.36%)** | **+146/6,717** | **+28/1,072** |

<sup>a</sup>Seven individuals without Test in cycle 1, but subsequent Test in cycle 2 and/or cycle 3.

The SARS-CoV-2 serology assay used in the initial phase of this study was a lab developed test based on the Epitope Diagnostics Coronavirus (COVID-19 IgG ELISA Kit: (http://www.epitopediagnostics.com/covid-19-elisa). This assay utilizes a microplate-based enzyme immunoassay technique. Assay controls and 1:100 diluted human serum samples were added to the microtiter wells of a microplate that was coated with SARS-CoV-2 recombinant full length nucleocapsid protein. After the first incubation period, the unbound protein matrix was removed with a subsequent washing step. A horseradish peroxidase (HRP) labeled polyclonal goat anti-human IgG tracer antibody was added to each well. After an incubation period, an immunocomplex of SARS-CoV-2 recombinant antigen – human anti-SARS-CoV-2 IgG antibody – HRP labeled antihuman IgG tracer antibody is formed if there is specific coronavirus IgG antibody present in the tested specimen. The unbound tracer antibody was removed by the subsequent washing step. HRP tracer antibody bound to the well is then incubated with a substrate solution in a timed reaction and then measured in a spectrophotometric microplate reader. The enzymatic activity of the tracer antibody bound to the anti-SARS-CoV-2 IgG on the wall of the microtiter well is proportional to the amount of the anti-SARS-CoV-2 IgG antibody level in the tested specimen. Qualitative results and optical densities (OD) values reported by the instrument were used in the analysis. The assay used in the initial phase of this study was validated by the ELISA vendor in a cohort of 84 patients from Jiaxing City Center for Disease Control and Prevention and Zhejiang University Hospital, with a reported sensitivity and specificity of 100%. Subsequently, the assay was internally validated as a laboratory developed test using a second cohort of 91 patients, collected at Providence Portland Medical Center, and including RT-PCR confirmed positive hospitalized patients and in sera collected in 2019 before the SARS-CoV-2 initial outbreak. In this cohort, the sensitivity was 80%, the specificity 100%, which is in line with another recent external performance evaluation^14^. Possible limitations of this assay include cross-reactivity due to past or present infection with non-SARS-CoV-2 coronavirus strains. Sensitivity and specificity have not been established in the setting of asymptomatic SARS-CoV-2 infection.

Odds ratios and two-sided CIs for seropositivity risk by sex, age (< 50 / 50+), role, hospital, and workplace were determined via logistic regressions comparing each group against all others in that category (e.g., physicians versus all other job roles combined) with age and sex as covariates. Cohen’s kappa with two-sided asymptotic CIs and positive and negative percent agreement were used to assess concordance between swab qPCR and serological results^15–17^. No data were imputed. Analyses were performed in R version 3.6.2, release 2019–12–12 (see supplemental for code)^18^.

## RESULTS

The pace of HCW voluntary enrollment was brisk, with a single-day maximum of 1,293 phlebotomies performed. In total, 10,019 asymptomatic HCW took part in the study, with blood draws every 14 +/− 3 days over a four week interval (up to three blood draws), at eight hospital medical centers across the state of Oregon, USA. Participants were 75.74% female (7,588/10,019), with a median age of 42 years (range 18 to 82).

The overall 4-week cross-sectional seroprevalence was 2.53% (+253/10,019). Detailed testing results are presented in Table 1, including the specific range of testing dates at each hospital medical center. Figure 1 presents serosurveillance relative to aggregate daily COVID-19 hospital census at the participating Oregon medical centers (for breakdown by individual hospital site, see supplemental Figure S1).

**Figure 1.**
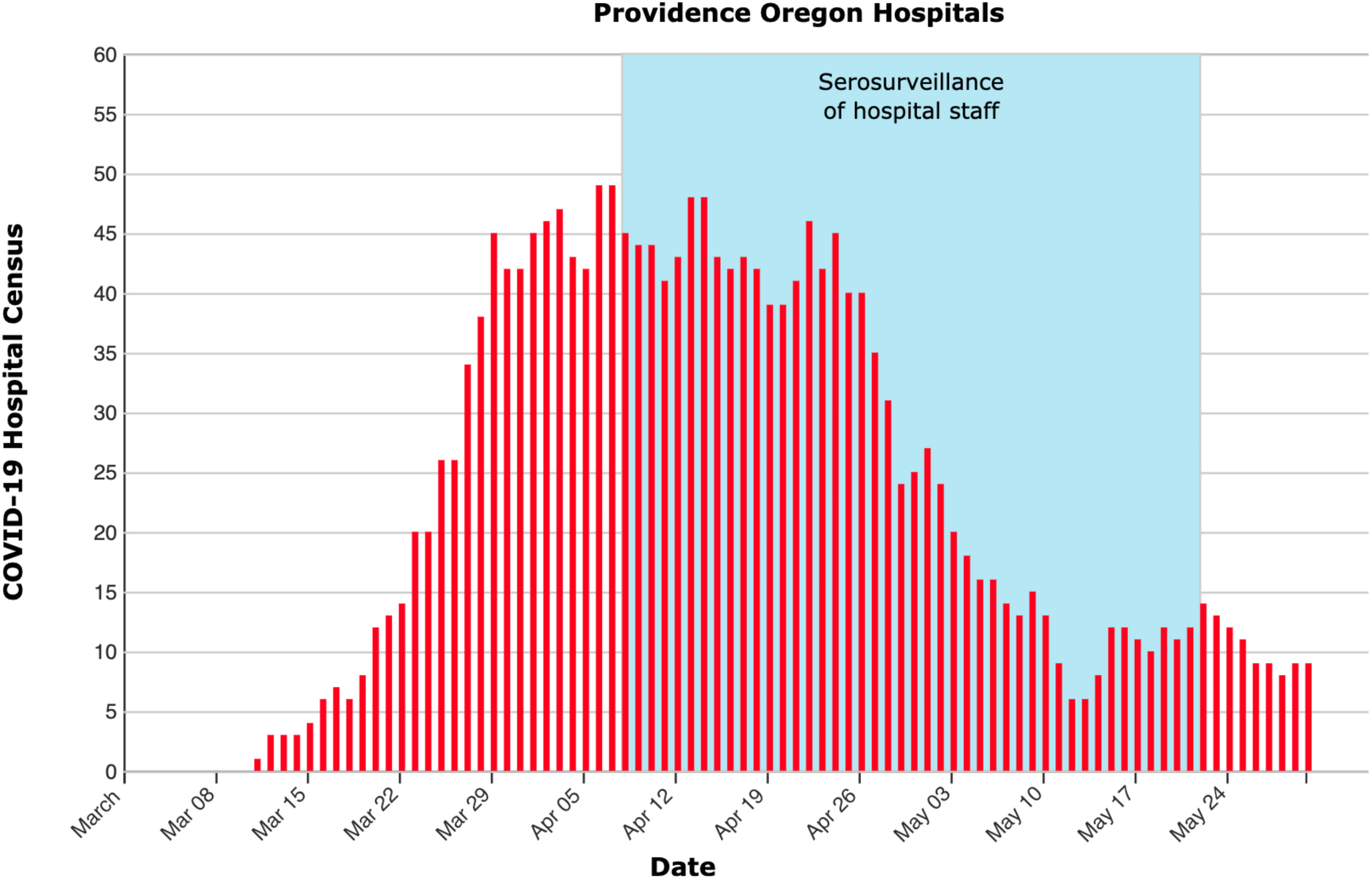
Serosurveillance Providence Oregon Cohort Asymptomatic Health Care Workers

As shown in Figure 2, we observed significantly increased seropositivity among HCW age 50 and above, with odds ratio of 1.51 (95% CI 1.17–1.94). We also observed geographic variation in HCW seroprevalence based on urban versus non-urban regions, ranging from 3.04% in the Portland, Oregon metropolitan area, to 1.36% in non-urban medical centers, which correlated with the overall geographic distribution of COVID-19 cases in the state during the same period.

**Figure 2.**
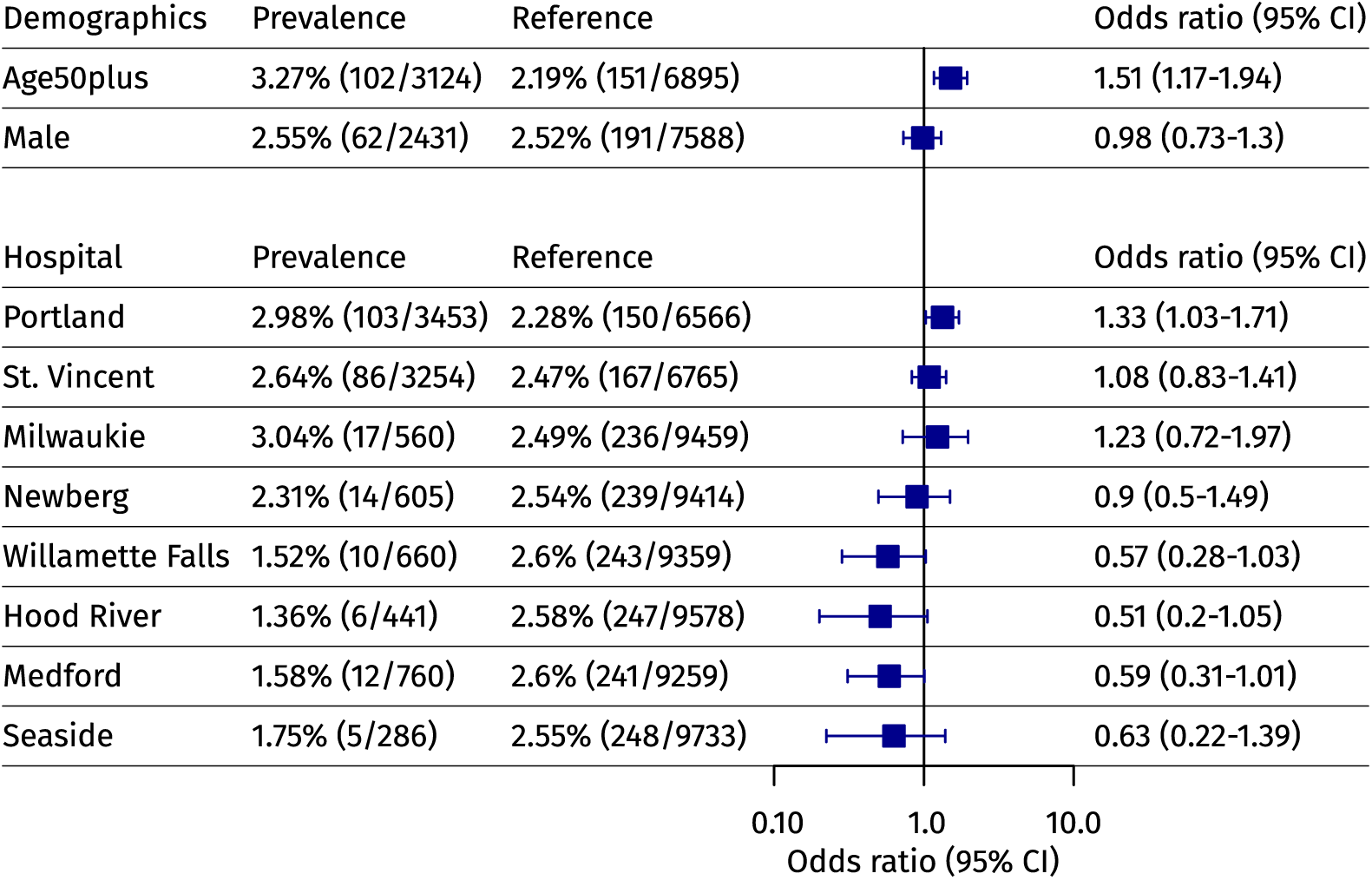
Demographic and Geographic Variation, Odds Ratios (95% CI)

As shown in Figure 3, subgroup analysis by HCW job role identified significant differential seroprevalence. Housekeepers were at the most significantly increased risk of seropositivity, with 8.03% seroprevalence (+11/137) and an odds ratio of 3.17 (95% CI 1.59–5.71); whereas, anesthesiologists were at the most significantly decreased risk of seropositivity, with 0.00% seroprevalence (0/110) and an odds ratio of 0.00 (95% CI 0–0.26). Subgroup analysis by workplace setting, however, did not identify significant differential seroprevalence, as shown in Figure 4. For a complete listing of subgroup classifiers and results, see supplemental Tables S1 and S2.

**Figure 3.**
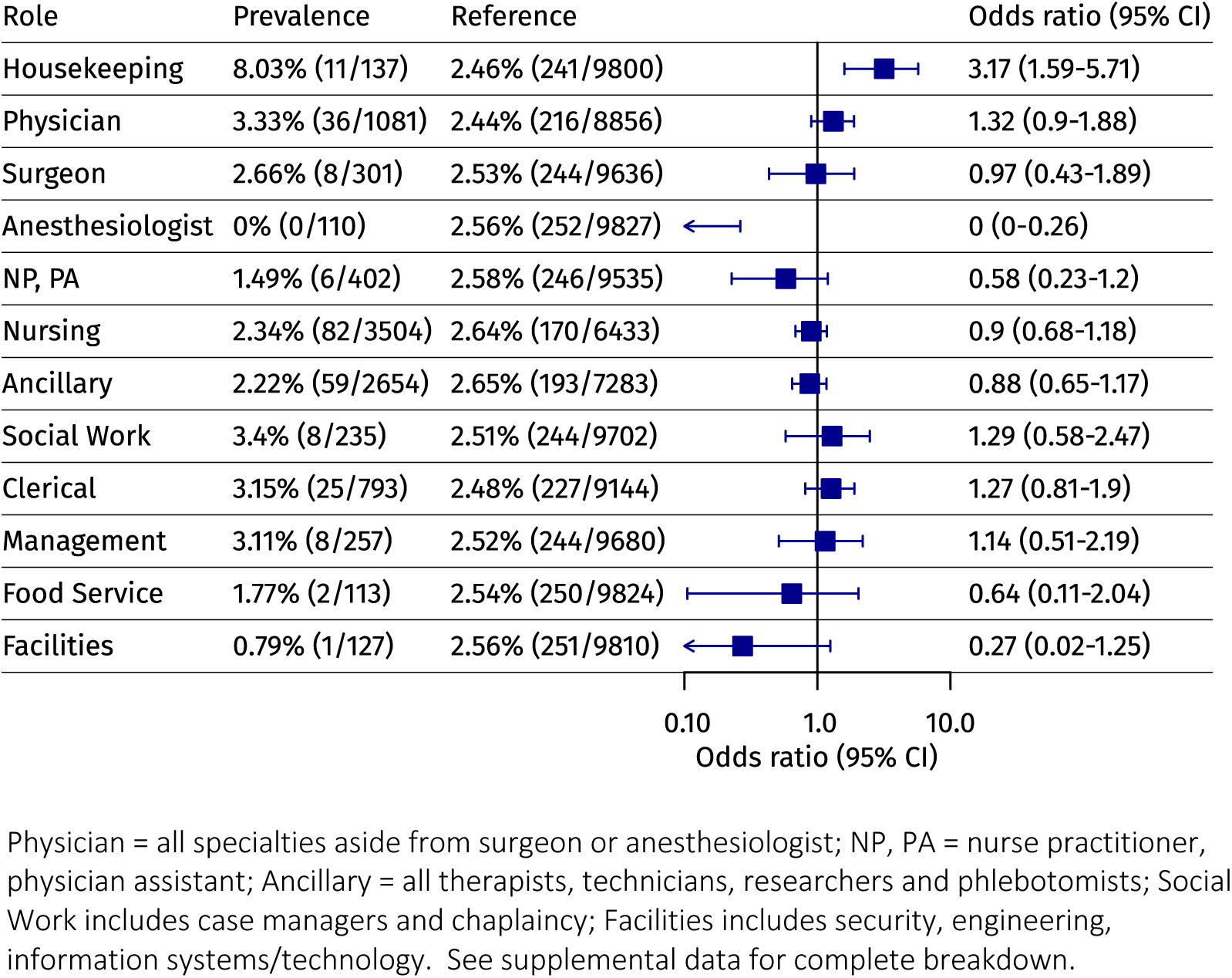
Job Role Subgroup Analysis, Odds Ratios (95% CI)

**Figure 4.**
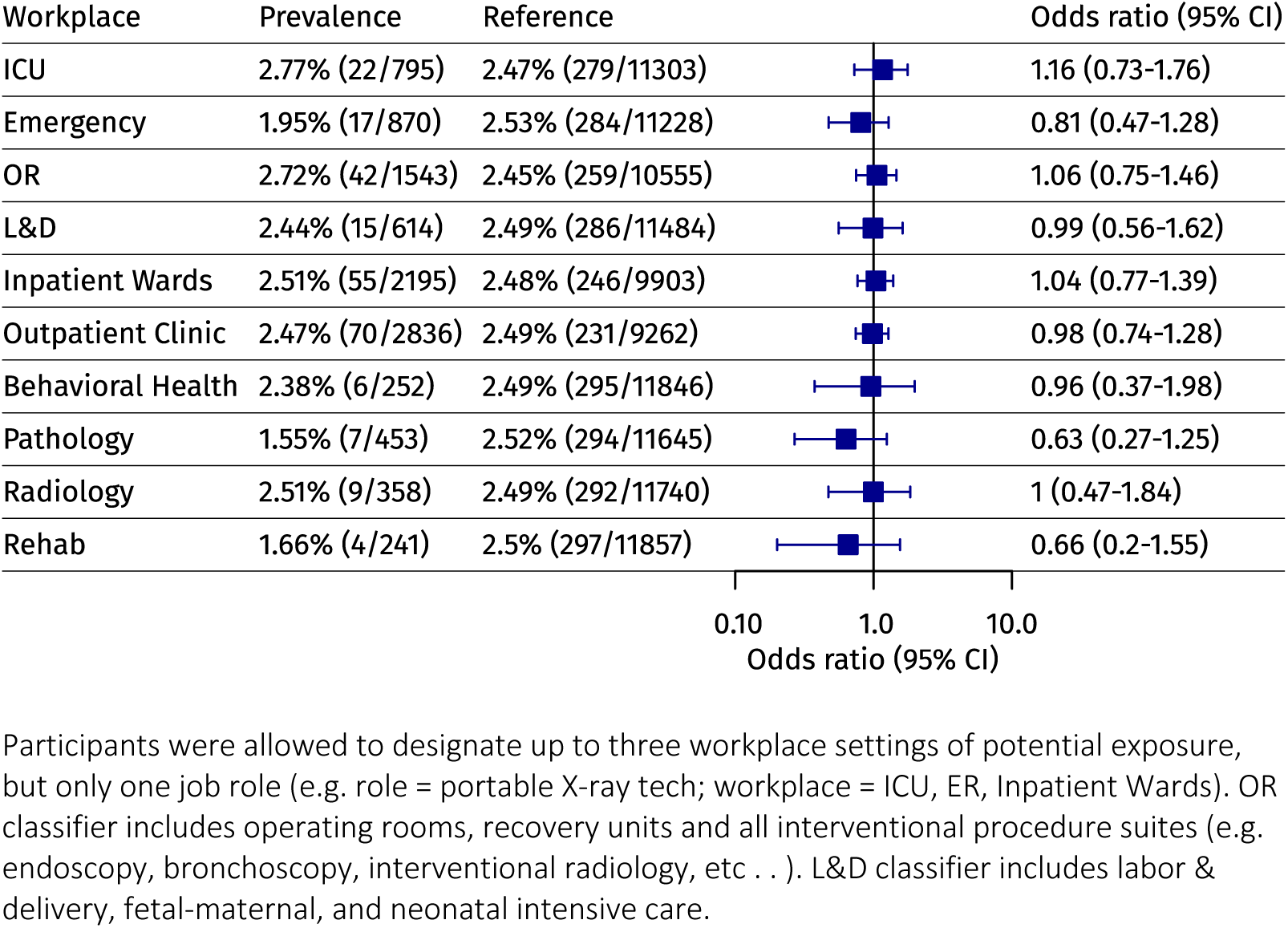
Workplace Subgroup Analysis, Odds Ratios (95% CI)

Longitudinal serosurveillance over the course of the study, as shown in Table 2, identified 17 seroconversion events, (0.25%; 17 of 6,717), and 101 seroreversion events (1.50%; 101 of 6,717). As shown in Table 3, prior qPCR-based SARS-CoV-2 swab testing results were self-reported by 1,048 study participants: of 1,021 negative qPCR swab tests reported, 995 were also negative by serology (97.45% negative agreement); while, of 27 positive qPCR swab tests, 17 were also positive by serology (62.96% positive agreement); **κ** = 0.47 (95% CI 0.32–0.62).

**Table 2.** Seroconversion and Seroreversion Events

| Providence Medical Center | Ever-Positive/Total (%) | Seroconversion | Seroreversion |
| --- | --- | --- | --- |
| Portland | +103/3,453 (2.98%) | 9 | 57 |
| St. Vincent | +86/3,254 (2.64%) | 7 | 25 |
| Milwaukie | +17/560 (3.04%) | 0 | 6 |
| Newberg | +10/660 (1.52%) | 0 | 3 |
| Willamette Falls | +14/605 (2.31%) | 0 | 2 |
| Hood River | +6/441 (1.36%) | 1 | 2 |
| Medford | +12/760 (1.58%) | 0 | 5 |
| Seaside | +5/286 (1.75%) | 0 | 1 |
| **Totals** | **+253/10,019 (2.53%)** | **+17/6,717 (0.25%)** | **101/6,717 (1.50%)** |

**Table 3.** Swab qPCR versus ELISA Serology

| SARS-CoV-2 swab qPCR (1,048) | IgG nucleocapsid ELISA |  | % Agreement |
| --- | --- | --- | --- |
|  | Positive | Negative |  |
| Negative (1,021) | +26 | 995 | 97.45% |
| Positive (+27) | +17 | 10 | 62.96% |

## DISCUSSION

On Wednesday, April 1, 2020, Dr. Deborah Birx, the White House coronavirus coordinator, issued a call to action to academia to develop tests quickly for health care worker serosurveillance. “We have the most brilliant scientists in the world in our universities in state after state. Our universities can do that by Friday. So I’m putting that challenge out to them to really work on that and do that… they could screen the entire hospital.”^19^ Hence, the Providence Oregon HCW serosurveillance effort was operationalized in one week (April 1 to 8, 2020), from study conception to protocol writing, to IRB approval, to administrative review, to secure intranet architecture build for results notification, to staffing at scale, to logistics, to on-site training, to first blood draw.

At the time, little data existed as to asymptomatic seroprevalence among HCW or other groups. Preliminary data from two population serosurveys were reported in the press. In San Miguel County, Colorado USA, a program to offer county residents free and voluntary antibody testing was launched in Telluride on March 24^th^ 2020, and as of early April, a 0.5% cross-sectional seroprevalence emerged (+8 of 1,631)^20,21^. Also in early April, researchers from the University Hospital in Bonn, Germany released preliminary findings of a serosurvey in Gangelt, the epicenter of a point outbreak during Karneval celebrations a month earlier, showing 14% seroprevalence and a 2% rate of residual active infection by tandem swab qPCR.^22,23^

Results are presented here from a SARS-CoV-2 serosurveillance study in a cohort of 10,019 asymptomatic health care workers (HCW) in Oregon, USA, as part of a larger serology screening effort that is being extended to approximately 100,000 caregivers employed by a large integrated not-for-profit health care system spanning seven western states (Providence St. Joseph Health, PSJH). Serosurveillance over a 4-week time period in April-May 2020, during the peak of initial COVID-19 hospital case load, was free & voluntary with available blood draws and confidential delivery of results to participants every 14 +/− 3 days. The first site was activated on April 8^th^ 2020 at a large urban Portland medical center, followed by a staggered roll-out to seven additional PSJH hospital centers across the state of Oregon, with closure of the final site on May 22^nd^ 2020. Antibody screening was performed via a lab developed test ELISA based on the Epitope Diagnostics Coronavirus COVID-19 IgG nucleocapsid detection Kit^14^. Overall, we identified 253 SARS-COV2 IgG seropositive individuals, representing a cross-sectional seroprevalence of 2.53% for asymptomatic HCWs, in the 10,019 participant Providence Oregon cohort.

Given the low seroprevalence levels we observed, it is notable that significant differences among subgroups emerged nonetheless. Seropositivity was most significantly increased among housekeeping staff (odds radio 3.17; 95% CI 1.59–5.71) and in general for asymptomatic HCW age 50 and above (odds ratio 1.51, 95% CI 1.17–1.94); whereas, to our surprise, seropositivity was most significantly decreased among anesthesiologists (odds ratio 0.00; 95% CI 0.00–0.26). The identification of anesthesiologists as the occupational subgroup with the most reduced risk of seropositivity in this large multi-hospital cohort was unexpected, given specific occupational exposure risks inherent to this specialty, and likely attests to the strong regimes of prevention and protection instituted in the work environment. However, equally noteworthy is the identification of housekeeping staff at the opposite pole of risk-extreme, particularly, as this finding directly parallels the recent HCW data reported by *Shields et al*. at University Hospital Birmingham, UK^24^. While raising the possibility of differential occupational exposure, these findings may also represent the effect of underlying socioeconomic disparities among HCW subgroups.

Longitudinal serosurveillance over a four-week window, identified 17 seroconversion events (0.25%) across the eight participating hospital sites, with a substantially higher rate of 101 seroreversion events (1.50%). The very low level of seroconversion suggests that rigorous enforcement of PPE and of statewide aggressive social distancing measures may have been major determinants in minimizing SARS-CoV-2 infection rates among HCW. On the other hand, the relatively higher rates of seroreversion, may be in line with recent reports of COVID-19 antibody titers declining rapidly to non-detectable levels following mild or asymptomatic disease^9–12^. We also observed relatively poor positive agreement (63%) between serology and self-reported positive swab qPCR; whereas, negative agreement between serology and self-reported negative swab qPCR was relatively high (97%). Although exact dates of prior qPCR swab testing were not captured, this finding is also likely supported by recent reports of antibody titer decline^9–12^, notwithstanding the fact that we assessed only SARS-CoV-2 nucleocapsid IgG, and the possibility that other SARS-CoV-2 antigens may ultimately be shown to be more reliable and long-lasting serologic targets.

Overall, our findings indicate relatively low seroprevalence and very low seroconversion rates among HCW during the initial surge of COVID-19 hospital case load in Oregon, USA, a period in which aggressive social distancing measures were in place. The high seroreversion rate and the relatively high discordances between SARS-CoV2 serology and swab qPCR assessment (**κ** = 0.47; 95% CI 0.32–0.62), also highlight limitations of current detection methods and stress the need for development of novel assessment methodologies. Use of residual sera is permitted in the study for further immunologic analyses, such as orthogonal serologic assay methods. It remains unclear whether the humoral antibody response is fully protective, or whether cellular immunity may be playing a larger role in the clearance of SARS-CoV-2^13,25,26^. In the event of future SARS-CoV-2 seasonal cycling, this protocol allows for reactivation without formal re-review in order to enable timely data capture on re-emergent pandemic/endemic conditions.

## Data Availability

All data available in supplemental section.

## REFERENCES

1. Cucinotta, D. & Vanelli, M. WHO Declares COVID-19 a Pandemic. Acta Bio Medica Atenei Parmensis 91, 157–160 (2020).

2. Grasselli, G., Pesenti, A. & Cecconi, M. Critical Care Utilization for the COVID-19 Outbreak in Lombardy, Italy: Early Experience and Forecast During an Emergency Response. JAMA 323, 1545–1546 (2020).

3. Lewnard, J. A. & Lo, N. C. Scientific and ethical basis for social-distancing interventions against COVID-19. Lancet Infect Dis 20, 631–633 (2020).

4. The Lancet. COVID-19: protecting health-care workers. Lancet 395, 922 (2020).

5. Chirico, F., Nucera, G. & Magnavita, N. COVID-19: Protecting Healthcare Workers is a priority. Infection Control & Hospital Epidemiology 1–1 (undefined/ed) doi:10.1017/ice.2020.148.

6. Zhan, M., Qin, Y., Xue, X. & Zhu, S. Death from Covid-19 of 23 Health Care Workers in China. New England Journal of Medicine 382, 2267–2268 (2020).

7. Holshue, M. L. et al. First Case of 2019 Novel Coronavirus in the United States. N Engl J Med 382, 929–936 (2020).

8. Chou, R. et al. Update Alert 2: Epidemiology of and Risk Factors for Coronavirus Infection in Health Care Workers. Ann. Intern. Med. (2020) doi:10.7326/M20-4806.

9. Long, Q.-X. et al. Clinical and immunological assessment of asymptomatic SARS-CoV-2 infections. Nature Medicine 1–5 (2020) doi:10.1038/s41591-020-0965-6.

10. Liu, T. et al. Prevalence of IgG antibodies to SARS-CoV-2 in Wuhan – implications for the ability to produce long-lasting protective antibodies against SARS-CoV-2. medRxiv 2020.06.13.20130252 (2020) doi:10.1101/2020.06.13.20130252.

11. Ibarrondo, F. J. et al. Rapid Decay of Anti–SARS-CoV-2 Antibodies in Persons with Mild Covid-19. New England Journal of Medicine 0, null (2020).

12. Seow, J. et al. Longitudinal evaluation and decline of antibody responses in SARS-CoV-2 infection. medRxiv 2020.07.09.20148429 (2020) doi:10.1101/2020.07.09.20148429.

13. Yongchen, Z. et al. Different longitudinal patterns of nucleic acid and serology testing results based on disease severity of COVID-19 patients. Emerging Microbes & Infections 9, 833–836 (2020).

14. Whitman, J. D. et al. Test performance evaluation of SARS-CoV-2 serological assays. medRxiv (2020) doi:10.1101/2020.04.25.20074856.

15. McHugh, M. L. Interrater reliability: the kappa statistic. Biochem Med (Zagreb) 22, 276–282 (2012).

16. Cohen, J. A coefficient of agreement for nominal scales. Educational and Psychological Measurement 20, 37–46 (1960).

17. Jørgensen, J. T. et al. High Concordance Between Two Companion Diagnostics TestsA Concordance Study Between the HercepTest and the HER2 FISH pharmDx Kit. Am J Clin Pathol 136, 145–151 (2011).

18. Meier, K. Guidance for Industry and FDA Staff – Statistical Guidance on Reporting Results from Studies Evaluating Diagnostic Tests. 39.

19. Create Antibody Tests for Health Workers, Birx Urges Researchers (1). https://news.bloomberglaw.com/pharma-and-life-sciences/create-antibody-tests-for-health-workers-birx-urges-researchers.

20. Zhang, S. Why a Tiny Colorado County Can Offer COVID-19 Tests to Every Resident. The Atlantic https://www.theatlantic.com/science/archive/2020/03/coronavirus-tests-everyonetiny-colorado-county/608590/ (2020).

21. Aschwanden, C. Kaiser Health News – A Colorado Ski Community Planned To Test Everyone For COVID-19. Here’s What Happened. Kaiser Health News https://khn.org/news/a-colorado-ski-community-planned-to-test-everyone-for-covid-19-heres-what-happened/ (2020).

22. Blood tests show 14% of people are now immune to covid-19 in one town in Germany. MIT Technology Review https://www.technologyreview.com/2020/04/09/999015/blood-tests-show-15-of-people-are-now-immune-to-covid-19-in-one-town-in-germany/.

23. http://zwischenergebnis_covid19_case_study_gangelt_en.pdf.

24. Shields, A. M. et al. SARS-CoV-2 seroconversion in health care workers. medRxiv 2020.05.18.20105197 (2020) doi:10.1101/2020.05.18.20105197.

25. Wu, F. et al. Neutralizing antibody responses to SARS-CoV-2 in a COVID-19 recovered patient cohort and their implications. medRxiv 2020.03.30.20047365 (2020) doi:10.1101/2020.03.30.20047365.

26. Le Bert, N. et al. SARS-CoV-2-specific T cell immunity in cases of COVID-19 and SARS, and uninfected controls. Nature (2020) doi:10.1038/s41586-020-2550-z.

